# Longitudinal associations between perceived stress and anhedonia during psychotherapy

**DOI:** 10.1101/2022.03.09.22272139

**Authors:** Rachel Phillips, Erin Walsh, Todd Jensen, Gabriela Nagy, Jessica Kinard, Paul Cernasov, Moria Smoski, Gabriel Dichter

## Abstract

**Background:** Chronic stress alters reward sensitivity and contributes to the emergence of anhedonia. In clinical samples, the perception of stress is a strong predictor of anhedonia. While there is substantial evidence demonstrating psychotherapy reduces perceived stress, little is known regarding the effects of treatment-related decreases in perceived stress on anhedonia.

**Methods:** The current study investigated reciprocal relations between perceived stress and anhedonia using a cross-lagged panel model approach in a 15-week clinical trial examining the effects of Behavioral Activation Treatment for Anhedonia (BATA), a novel psychotherapy to treat anhedonia, compared to a Mindfulness-Based Cognitive Therapy (MBCT) comparison intervention (ClinicalTrials.gov Identifiers NCT02874534 and NCT04036136).

**Results:** Treatment completers (*n*=72) experienced significant reductions in anhedonia (*M*=-8.94, *SD*=5.66) on the Snaith-Hamilton Pleasure Scale (*t*(71)=13.39, *p*<.0001), and significant reductions in perceived stress (*M*=-3.71, *SD*=3.88) on the Perceived Stress Scale (*t*(71)=8.11, *p*<.0001) following treatment. Across all treatment-seeking participants (*n*=87), a longitudinal autoregressive cross-lagged model revealed significant paths showing that higher levels of perceived stress at treatment Week 1 predicted reductions in anhedonia at treatment Week 4.

**Conclusions:** Longitudinal models illustrated that individuals with relatively high perceived stress at the start of treatment were likely to report relatively lower anhedonia a few weeks into treatment. At mid-treatment, individuals with low perceived stress were more likely to report lower anhedonia towards the end of treatment. Early treatment components are thought to reduce perceived stress, allowing for mid-to-late treatment components to exert their direct effects on anhedonia. The findings presented here demonstrate the importance of including stress-reducing components in cognitive-behavioral-based anhedonia treatments.

## Introduction

Anhedonia, the loss of interest or reduced pleasure in activities that were previously rewarding (APA, 2013; Tolentino & Schmidt, 2018), is a hallmark symptom of numerous psychiatric disorders (Der-Avakian & Markou, 2012). There is strong evidence that chronic stress contributes to the emergence of anhedonia (Bogdan et al., 2012; Ironside et al., 2018; Kumar et al., 2015; Pelletier-Baldelli et al., 2021; Pizzagalli, 2014; Pizzagalli et al., 2007). In clinical samples, the perception of stress is a strong predictor of risk for psychopathology (Conway et al., 2012; Hammen et al., 1992; Kupferberg et al., 2016).

One model of the relationship between stress and psychopathology—including anhedonia—is the “kindling” or stress sensitization theory. This theory posits that the relationship between stress and psychopathology changes over time, such that psychiatric symptoms are initially precipitated by a major stressful life event and recurrent symptoms are triggered by less severe (i.e., minor) stressors. This process of sensitization--or increased sensitivity to subsequent, minor stressors—is thought to confer greater risk for psychiatric symptoms and disorders over time. Consistent with this theory, research shows that for adults with psychiatric conditions where anhedonia is centrally relevant (e.g., depression), the onset of disorder is strongly associated with severe life events and recurrences are associated with lower severity events (Kendler et al., 2003; Stroud et al., 2011). Although the severity of lifetime stressors is important, the perception of stressful events is also critical for understanding the etiology and recurrence of psychopathology (Hewitt et al., 1992).

Perceived stress can be considered the subjective significance of stressors and defined as “the degree to which individuals appraise situations in their lives as stressful” (Hewitt et al., 1992). As an approach to stress measurement, perceived stress contrasts objective stress measures such as frequency of major life events; yet is nonetheless important for understanding individual differences in coping (Kanner et al., 1981; Lazarus & Folkman, 1986). When stressful life events are perceived or interpreted to have negative implications for one’s future or for their own self-worth, cognitively vulnerable individuals are susceptible to negative outcomes (Haeffel et al., 2008). A perceived lack of control and unpredictability of stressors is linked to a depressive attributional style, wherein there is a tendency to attribute stressors to causes that are “internally located, stable in time, and global in scope” (Willner et al., 1990). Furthermore, perceptions of stress as uncontrollable and unpredictable predict reduced goal-directed behavior (Haeffel et al., 2008; Pizzagalli, 2014). Specifically, perceived lack of control over stressors reduces reward responsiveness (Abramson et al., 1978; Pizzagalli, 2014), which in turn may predict anhedonia or be associated with compensatory reward-seeking behavior (Pechtel et al., 2013; Pechtel & Pizzagalli, 2013).

Across psychotherapy and pharmacotherapy mood disorders treatment studies, perceived stress is shown to decline alongside depressive symptoms (Conklin et al., 2020; A. Farabaugh et al., 2015). A positive affect treatment for depression and anxiety, for example, improved positive affect while lowering self-reported stress, depression, and anxiety in individuals with clinically-elevated symptoms (Craske et al., 2019). Moreover, there is evidence for reductions in anhedonia following pharmacotherapies (e.g., oxytocin) that increase motivational salience of rewards and reduce psychosocial stress reactivity (Sippel et al., 2017). Antidepressants exert clinical effects by helping to reduce anhedonia, following uncontrollable and unpredictable stressors, as demonstrated in animal models of anhedonia (Casarotto & Andreatini, 2007; Muscat et al., 1992; Willner et al., 1994). There remains a gap in understanding changes in anhedonia and stress during psychotherapy, not specific to depression. The current study aims to bridge this gap in recent work, which has been limited to human psychotherapeutic trials focused on reducing depressive symptoms on one side, and pharmacotherapy trials in animals that have examined treatment related changes in stress-induced anhedonia on the other.

The purpose of this study was to evaluate relations between concurrent changes in self-reported anhedonia and perceived stress over the course of psychotherapy for anhedonia. While it is well established that lower levels of reported stress are associated with fewer depressive symptoms (Hammen, 2005; Pegg et al., 2019) and particularly reduced anhedonia (Bogdan et al., 2012; Pelletier-Baldelli et al., 2021; Pizzagalli, 2014), this study examined whether differences in perceived stress precede or follow changes in anhedonia severity over the course of therapy. To investigate these treatment-related changes, this study examined self-reported anhedonia and stress from an ongoing clinical trial evaluating a novel psychotherapy, Behavioral Activation Treatment for Anhedonia (BATA), relative to a Mindfulness-Based Cognitive Therapy (MBCT) comparison intervention, for individuals with clinically elevated transdiagnostic anhedonia.

BATA is a modification of Behavioral Activation Treatment for Depression (Lejuez et al., 2011) and was developed to treat anhedonia regardless of clinical diagnosis. As in traditional behavioral activation approaches, BATA includes elements of psychoeducation, behavioral monitoring, and values assessment. The therapist supports patients in developing, setting, and achieving values-congruent behavioral goals. While achievable goal-setting is often impaired in anhedonia due to deficits in effort valuation and reward-based learning, BATA therapists provide structure and support in goal-setting and overcoming behavioral avoidance. In addition to traditional behavioral activation elements, BATA includes modules that focus on initiating new behaviors outside the established behavioral set (i.e., “dabbling”) and moment-savoring exercises to enhance consummatory reward experiences. Increased positive affect and decreased negative affect are theorized to result from reduced behavioral avoidance and consequently increased contact with potential reinforcers.

MBCT is traditionally administered in a group format and includes therapist-guided in-session meditation practices, inquiry of experience during practice as well as differences between mindfulness and everyday patterns of thought and behavior (Segal et al., 2018). MBCT was modified in the current study to an individual format, consistent with (Schroevers et al., 2015). Mindfulness is presented as a means to facilitate flexible cognitive-emotional responses to events and reduce habitual reactions. Practices emphasize core mindfulness skills of attention, decentering, and nonjudgmental acceptance through in-session and home practice exercises such as body scans, mindful movement, and focused awareness of breath. Guided inquiry is geared toward awareness of the interrelations among thoughts, emotions, and sensations, as well as the recognition of habitual patterns that can interfere with the experience of pleasure. While neither treatment targets stress reduction directly, the skills taught in MBCT have been effectively shown to reduce perceived stress (Alsubaie et al., 2017).

In the present study, we examined the longitudinal relations between perceived stress and anhedonia not specific to the effects of BATA vs. MBCT. Interim results from this trial suggest no difference in impact of the two treatments on anhedonia symptoms (Cernasov et al., 2021). Across both treatments, we hypothesized that psychotherapy treatment would lead to reductions in (a) perceived stress, and (b) anhedonia. Next, because it has been shown that stress alters reward sensitivity and contributes to the emergence of anhedonia (Kujawa et al., 2020; Kumar et al., 2015), we expected that treatment reductions in perceived stress would be significantly associated with treatment reductions in anhedonia.

Using longitudinal modeling approaches, we then explored reciprocal relations between perceived stress and anhedonia over time. We hypothesized cross-lagged effects (i.e., from one variable to another across time) of perceived stress on anhedonia, such that individuals with lower perceived stress would experience subsequent decreases in anhedonia over the course of treatment, compared to individuals with higher perceived stress. Relatedly, we hypothesized that individuals with relatively lower perceived stress during treatment would experience a subsequent decrease in anhedonia. Finally, we hypothesized that perceived stress would be inversely related to anhedonia, such that reductions in anhedonia symptoms following psychotherapy treatment would be dependent upon earlier levels of perceived stress. Overall, we expected these longitudinal models to reveal that changes in perceived stress predict changes in anhedonia across both treatments (Zyphur et al., 2020).

## Methods

### Participants

We analyzed data from an ongoing clinical trial for anhedonia that is evaluating the effectiveness of Behavioral Activation Treatment for Anhedonia (BATA) compared to Mindfulness-Based Cognitive Therapy (MBCT) in a transdiagnostic anhedonic sample (ClinicalTrials.gov Identifiers NCT02874534 and NCT04036136). The primary trial clinical endpoint is change in self-reported anhedonia. Eligible participants were 18 to 50 years old, actively seeking treatment for clinically significant anhedonia, measured by the Snaith-Hamilton Pleasure Scale (SHAPS; i.e., greater than or equal to 20; Franken et al., 2007; Pizzagalli et al., 2020), and met criteria for clinical impairment on the Clinician’s Global Impression Scale Severity (CGI-S; i.e., greater than or equal to 3).

Eighty-seven participants enrolled in the trial prior to October 2021 were randomly assigned to receive BATA (*n*=41) or MBCT (*n*=46). Participants completed up to 15 weeks of psychotherapy treatment for transdiagnostic anhedonia. If both therapist and participant agreed that maximum gains had already been reached after at least 8 sessions, treatment could be completed before 15 weeks. A proportion of patients (*n*=15) withdrew their participation prior to treatment completion. Participants enrolled after March 2020 received psychotherapy remotely, in response to the COVID-19 pandemic. All participants (*n*=87) were utilized for the longitudinal SEM models described below, while the subset of treatment completers (*n*=72) with available pre- and post-treatment scores on the Perceived Stress Scale (PSS) and SHAPS were included in preliminary regression analyses described below. Treatment completers include those who participated in a study completion visit, after completion of the recommended length of psychotherapy. Overall, participants completed an average of 11.24 weeks (*SD*=4.2) of psychotherapy treatment.

### Measures

The SHAPS is a well-validated 14-item questionnaire that assesses self-reported anhedonia. It was used to determine study eligibility (i.e., a score greater than or equal to 20) and to assess treatment response. A score of ≥ 20 corresponds to clinically significant anhedonia from a general population sample (Franken et al., 2007). Total scores on the SHAPS range from 14 to 56, whereby higher scores indicate greater anhedonia severity. The PSS is a widely used measure of perceived stress that is strongly associated with mental health outcomes (A. H. Farabaugh et al., 2004; Renwick et al., 2009). The PSS contains 10 items and assesses the extent to which an individual appraises nonspecific stressors as unpredictable and uncontrollable over the past month. Items are rated on a 1 (never) to 4 (very often; Hewitt et al., 1992). Total scores on the PSS range from 0 to 40, whereby higher scores indicate greater perceived stress. The PSS and SHAPS were collected at study assessments that occurred at baseline (Week 1) and Weeks 4, 8, 12, and 15 of psychotherapy treatment. Assessments could be conducted up to 1 week late (e.g., the Week 12 assessment could occur in week 12 or 13) to accommodate participant schedules.

### Statistical Analysis

Descriptive statistical analysis for all variables was conducted using R/RStudio (version 1.3.1093). Preliminary independent samples t-tests were conducted to determine whether treatment groups differed at baseline, Treatment Weeks 4, 8, 12, or 15 on anhedonia severity (i.e., SHAPS) or perceived stress (i.e., PSS). These assessments had a 2-week window for completion, so PSS and SHAPS data may have been collected +1 or -1 week from these benchmarks.

The current study evaluated treatment-related changes using multiple linear regression and structural equation modeling approaches. Multiple linear regressions controlling for sex, age, and treatment group (i.e., BATA, MBCT) examined relations between treatment-related change in anhedonia severity on the SHAPS and self-reported perceived stress on the PSS. Treatment-related changes on the PSS and SHAPS were calculated by subtracting pre-treatment scores from post-treatment scores, irrespective of the number of weeks in treatment. That is, the regression approach used here did not differentiate those who completed 8, 12, or 15 weeks of treatment. Analyses using a structural equation modeling framework were used to better understand change in symptoms at different weeks of treatment. These analyses were conducted using Mplus Version 8.4 (Muthén & Muthén, 2012). The SEM models described below (i.e., CLPM and RI-CLPM) allowed us to test the reciprocal relations among perceived stress and anhedonia longitudinally. Models were estimated using a maximum likelihood estimator, wherein full information maximum likelihood (FIML) was invoked for the context of handling missing data. FIML accounts for missing data by weighting cases with complete data more heavily and has been demonstrated to be more reliable than alternate approaches, including listwise deletion (Allison, 2003). Pre-specified indices used to evaluate model fit aligned with current standards in the field (Bowen & Guo, 2011; Kline, 2015) such that adequate fit between the model and the observed data were met if the (a) Chi-square (χ^2^) test had a *p*-value of ≥ 0.05, (b) Root mean square error of approximation (RMSEA) and the upper bound confidence interval were ≤ 0.08, and (c) Comparative fit index (CFI) and Tucker-Lewis index (TLI) were ≥ 0.95. While a non-significant (*p* ≥ 0.05) chi-square test is a relatively compelling indicator of model fit on its own, additional model fit indices are commonly used in the event of a significant chi-square test of model fit.

#### Cross-lagged Panel Model

Cross-lagged panel models (CLPM) are conceptually similar to traditional methods such as linear regression for longitudinal data. Both methods are used to evaluate relative change in the relationship between two constructs (Orth et al., 2021). However,7 a benefit of CLPM’s for longitudinal data is that they allow for multiple dependent variables, to test several directional relations at one time. An autoregressive cross-lagged panel model (CLPM) (**Fig 1**) was evaluated to test hypotheses. The following assumptions were followed: (a) anhedonia and stress were considered continuous variables, (b) perceived stress symptoms measured by the PSS and anhedonia symptoms measured by the SHAPS at the same timepoints were correlated (e.g., phi and psi coefficients), and (c) variables measured at different timepoints were associated with themselves longitudinally (e.g., auto-regressive paths). Standardized path coefficients were interpreted after controlling for time-invariant covariates of age, sex, and treatment group at treatment week 15.

**Fig 1.**
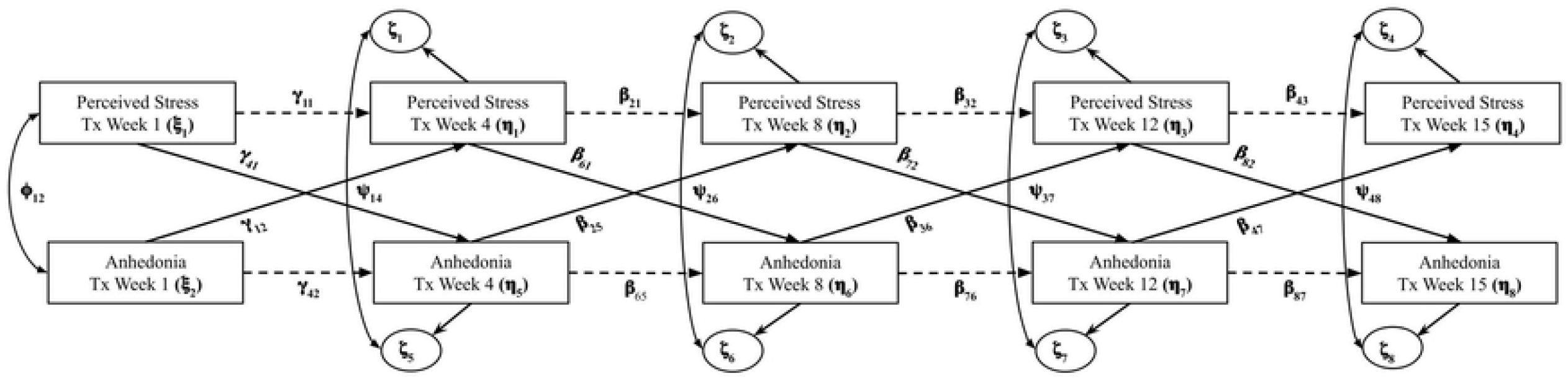
Autoregressive cross-lagged panel model of anhedonia and perceived stress over 15 weeks of psychotherapy treatment. Time-invariant covariates are age, sex, and treatment group, which were regressed on treatment (Tx) week 15 observed variables. Gamma coefficients represent the directional paths from exogenous to endogenous variables. Beta coefficients represent the directional paths between endogenous variables. Zeta coefficients represent the error variance terms for endogenous variables. Phi coefficients represent the covariances between exogenous variables. Psi coefficients represent the covariances between error variances linked to endogenous variables.

#### Random Intercept Cross-lagged Panel Model

While the CPLM is best suited for evaluating prospective effects of between-person differences, we also evaluated the prospective effects of within-person deviations using a random-intercept cross-lagged panel model (RI-CLPM) (Orth et al., 2021). The RI-CLPM has been introduced as an alternative CLPM that accounts for “stable, trait-like differences between units (e.g., individuals, dyads, families, etc.), such that the lagged relations pertain exclusively to within-unit fluctuations” (Mulder & Hamaker, 2020). Essentially, the RI-CLPM separates between-person variance (i.e., deviations in mean participant symptoms during treatment across the sample) from within-person variance (i.e., deviations among individual participant’s symptoms over treatment). In the RI-CLPM for this dataset, these cross-lagged paths represent within-person changes in symptoms during treatment. Auto-regressive effects represent the within-person stability of a particular domain (e.g., stress, anhedonia). Random intercepts (latent variables) in the model account for between-person differences (Orth et al., 2021).

## Results

### Sample Characteristics

The sample (*n*=87) ranged in age from 18 to 49 years (*M*=29.48, *SD*=9.07) and was comprised of 58 women and 29 men. Group differences between participants receiving BATA (*n*=41) and MBCT (*n*=46) were explored at baseline among measures of age, perceived stress, and anhedonia. No significant baseline differences between treatment groups were observed for age (*p*>.05), anhedonia on the SHAPS (*p*>.05), or perceived stress on the PSS (*p*>.05).

On average, participants across psychotherapy groups reported reduced perceived stress and anhedonia over the course of treatment (**Table 1**). There were significant reductions in SHAPS scores from Week 1 to Week 4 (*t*(75)=6.78, *p*<.001), Week 4 to Week 8 (*t*(61) = 5.18, *p* < .001), Week 8 to Week 12 (*t*(51) = 4.66, *p* < .001, and Week 12 to Week 15 (*t*(30) = 3.53, *p* = .0014). There were significant reductions in PSS scores from Week 4 to Week 8 (*t*(69) = 3.84, *p* < .001, and Week 12 to Week 15 (*t*(31) = 2.60, *p* = .014).

**Table 1.** Mean and standard deviation of PSS and SHAPS scores by time. Mean (SD).

| *Measure* | *Tx Week 1*<br>*n=86* | *Tx Week 4*<br>*n=84* | *Tx Week 8*<br>*n=70* | *Tx Week 12*<br>*n=54* | *Tx Week 15*<br>*n=36* |
| --- | --- | --- | --- | --- | --- |
| **Perceived Stress Scale (PSS)** | 21.74<br>(3.88) | 20.99<br>(3.38) | 19.39<br>(3.24) | 18.81<br>(4.60) | 17.50<br>(3.78) |
| *Measure* | *Tx Week 1*<br>*n=87* | *Tx Week 4*<br>*n=76* | *Tx Week 8*<br>*n=67* | *Tx Week 12*<br>*n=55* | *Tx Week 15*<br>*n=36* |
| **Snaith-Hamilton**<br>**Pleasure Scale (SHAPS)** | 37.34<br>(4.34) | 34.07<br>(4.73) | 30.78<br>(4.76) | 29.38<br>(5.71) | 28.11<br>(4.79) |

Treatment completers (*n*=72; 38 BATA, 34 MBCT), or those who completed the recommended length of psychotherapy, saw significant reductions in anhedonia symptoms (SHAPS: *M*=-8.94, *SD*=5.66) (*t*(71)=13.39, *p*<.0001), and significant reductions in perceived stress (PSS: *M*=-3.71, *SD*=3.88) (*t*(71)=8.11, *p*<.0001) from baseline to study completion. No significant differences between treatment groups (BATA, MBCT) were observed for change in SHAPS (*p*>.05) or PSS scores (*p*>.05).

### Associations Between Treatment-related Changes in Perceived Stress and Anhedonia

#### Multiple Linear Regression

The relationship between treatment-related declines in PSS and SHAPS were explored using linear regression. For treatment completers (*n*=72), reductions in SHAPS scores were significantly predicted by reductions in PSS scores (**Fig 2**) (β=.74, *t*=4.80, *p*<.001, _adj_R^2^=0.238). For each one point decrease in PSS scores over the course of treatment, there was a 0.74 unit decrease in SHAPS scores over the course of treatment. In this model, which controlled for sex, age, and psychotherapy treatment group (F=6.54, *p*<.001), reductions in PSS scores accounted for 23.8% of the variance in reductions in SHAPS scores. Interestingly, the reverse relationship was also observed, by which reductions in PSS scores were significantly predicted by reductions in SHAPS scores (β=.34, *t*=4.80, *p*<.001, _adj_R^2^=0.241). For each one point decrease in SHAPS scores, there was a 0.34 unit decrease in PSS scores over the course of treatment. In this model, which controlled for sex, age, and treatment group (F=6.62, *p*<.001), reductions in SHAPS scores accounted for 24.1% of the variance in reductions in PSS scores. Additionally, change on the SHAPS was positively correlated with change on the PSS (r=0.48). These results demonstrate that treatment-related reductions in anhedonia were accompanied by declines in self-reported perceived stress. However, this analytic approach alone cannot distinguish whether reductions in perceived stress precede or follow reductions in anhedonia.

**Fig 2.**
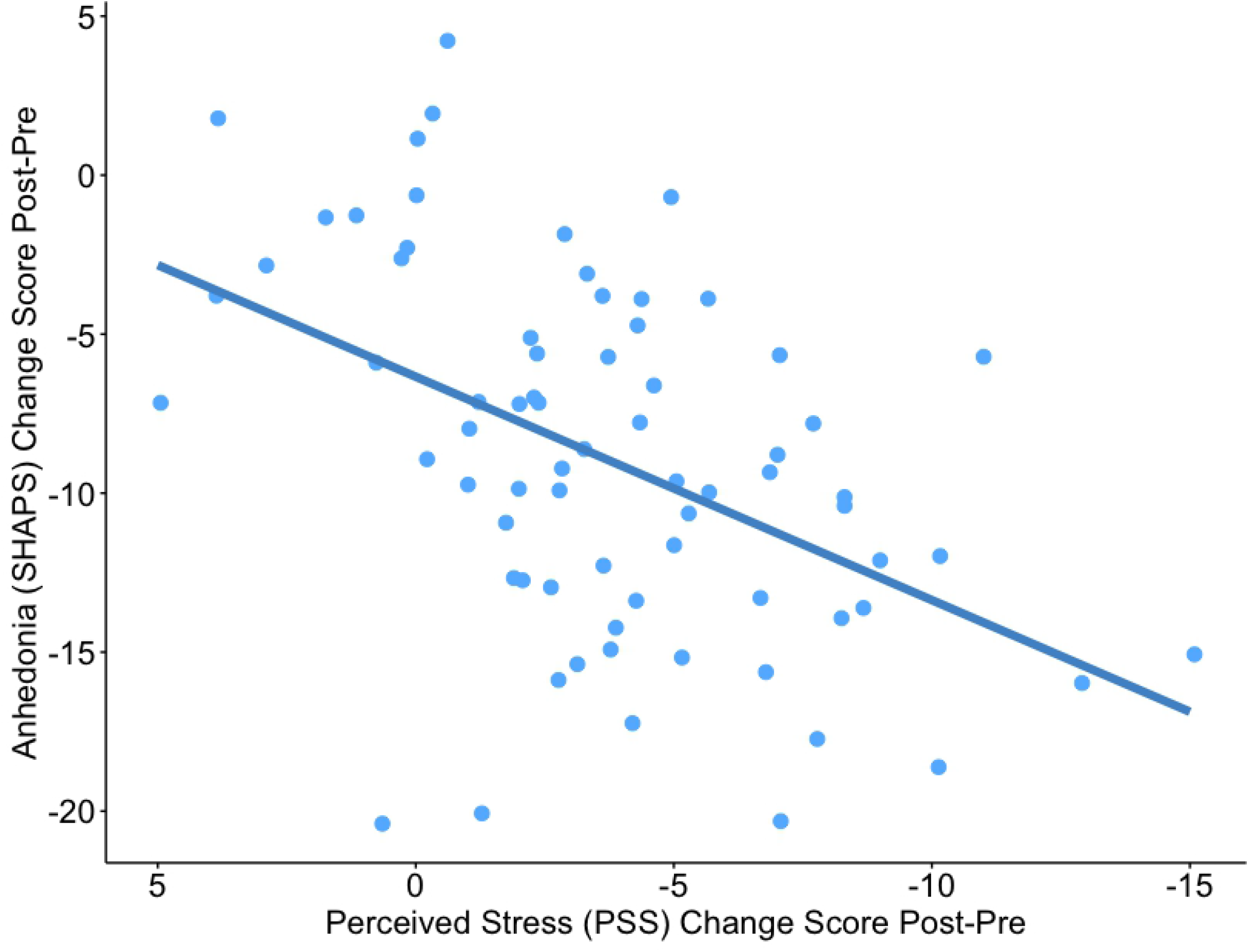
Treatment-related decline in perceived stress was associated with treatment-related decline in anhedonia severity. Multiple regression line for the association between change scores for perceived stress, measured by the Perceived Stress Scale (PSS), and change scores for anhedonia, measured by the Snaith-Hamilton Pleasure Scale (SHAPS). Change scores represent post-treatment minus pre-treatment scores. Covariates for age, sex, and treatment-group were included.

### Reciprocal Relations between Perceived Stress and Anhedonia at Different Treatment Stages

#### Cross-lagged Panel Model

To elucidate the degree to which changes in PSS scores were related to changes in SHAPS scores, a longitudinal structural equation model was evaluated. The autoregressive cross-lagged path model displayed in **Fig 3** met criteria for all pre-specified model fit indices except TLI and RMSEA upper bound: Chi-square χ^2^ (48) = 62.78, *p*=.074; Comparative fit index = 0.95; Tucker-Lewis index = 0.93; Root mean square error of approximation = 0.059 (90% CI = 0.000 – 0.097). For this reason, caution may be warranted in interpreting these findings, although taken together, the indices suggest adequate model fit. Significant positive autoregressive paths between perceived stress variables over time indicated the stability of this construct, as measured by the PSS. Positive autoregressive paths between anhedonia severity variables over time indicate the stability of anhedonia, as measured by the SHAPS. For both SHAPS and PSS scores, larger autoregressive coefficients closer to one reflect greater stability and less variance of these constructs over the course of treatment. One negative cross-lagged path from PSS scores at treatment Week 1 to SHAPS scores at treatment Week 4 emerged (β = -0.235, SE = 0.091, *p* = .010), indicating that PSS scores at the start of psychotherapy treatment significantly influenced SHAPS levels at treatment Week 4. A positive cross-lagged path from PSS scores at treatment week 8 to SHAPS at treatment week 12 also emerged (β = 0.193, SE = 0.074, *p* = .009). Detailed results are presented in **Supplementary Table 1**. There were no significant cross-lagged associations to support an effect of SHAPS scores on PSS scores at any stage of treatment.

**Fig 3.**
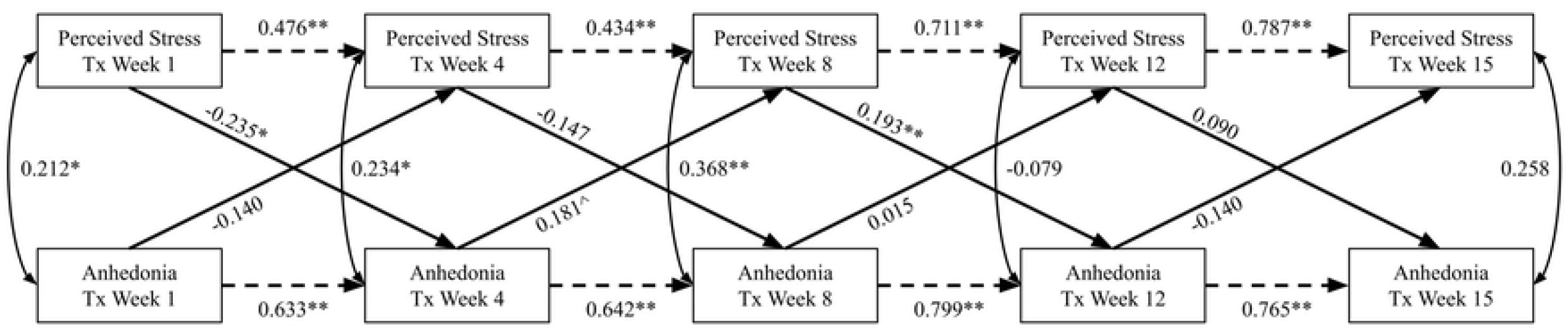
Estimated autoregressive cross-lagged model (CLPM) of anhedonia and perceived stress across psychotherapy treatment. One-headed dash lines represent auto-regressive paths; one-headed solid lines represent cross-lag paths; two-headed solid lines represent concurrent associations among variables at the same week in treatment. For ease of interpretation, error variance terms and covariates of age, sex, and treatment group are not included in the visualization. Values presented are standardized path coefficients.\*\**p*< 0.001, \**p*<.05, ^*p*<.10.

#### Random Intercept Cross-lagged Panel Model

The random-intercept autoregressive cross-lagged path model (RI-CLPM) displayed in **Supplementary Fig 1** showed poor fit: χ^2^ (54) = 112.8, *p*<.001; Comparative fit index = 0.79; Tucker-Lewis index = 0.71; root mean square error of approximation = 0.112 (90% CI = 0.083 – 0.141). This suggests that adequate fit between the RI-CLPM and the observed data was not met, and this model is uninterpretable.

## Discussion

The aim of the present study was to examine the relation between treatment-related changes in perceived stress and treatment-related changes anhedonia in an ongoing anhedonia clinical trial. We anticipated that individuals with clinically significant anhedonia would also report elevated perceived stress and hypothesized that anhedonia and perceived stress would be concurrently attenuated by psychotherapy.

First, we found that both psychotherapy treatments, Behavioral Activation Treatment for Anhedonia (BATA) and Mindfulness-based Cognitive Therapy (MBCT), effectively reduced anhedonia and perceived stress. There were no differences between treatment groups on baseline levels of anhedonia and perceived stress, nor changes in these measures by group over the course of treatment. Reductions in perceived stress and anhedonia are consistent with findings from previous randomized controlled trials that target positive and negative affect through both behavioral and mindfulness techniques (Colgan et al., 2019; Craske et al., 2019). These preliminary results allowed us to next examine the relation between perceived stress and anhedonia in the entire sample, regardless of psychotherapy received.

Across treatment groups, decreases in self-reported perceived stress were associated with decreases in self-reported anhedonia for treatment completers. For each one-point decrease in perceived stress over the course of treatment, there was a 0.74 unit decrease in anhedonia severity over the course of treatment. This supports known linkages between stress and anhedonia: Pizzagalli and colleagues (2007) found that individual differences in perceived stress are associated with reduced hedonic capacity, measured by reward responsiveness on a signal-detection task. As such, heightened perceived stress is likely to be associated with greater anhedonia. However, as presented above, we also found that treatment-related reductions in anhedonia significantly predicted reductions in perceived stress. These results indicated a bi-directional relationship between stress and anhedonia, so we proceeded with a prospective analysis of the reciprocal relation among perceived stress and anhedonia.

Through a longitudinal structural equation modeling approach, we found significant associations of perceived stress on subsequent levels of anhedonia. Results from a cross-lagged panel model (CLPM) showed significant effects of perceived stress at baseline on anhedonia at 4 weeks of psychotherapy treatment, as well as perceived stress at 8 weeks on anhedonia at 12 weeks of psychotherapy treatment. However, these effects differed in their direction, indicating a negative effect of perceived stress on anhedonia in early treatment and a positive effect of perceived stress on anhedonia in late treatment.

Early in psychotherapy treatment, cross-lagged associations indicated that higher levels of perceived stress at baseline (Week 1) were predictive of lower levels of anhedonia at Week 4. This association may be explained by early impact of therapeutic relationship and nonspecific therapy factors, including validation of the experience of anhedonia in both treatments. Both psychotherapy treatments administered here (i.e., BATA and MBCT) utilize psychoeducation about anhedonia and present a treatment model in early sessions, which may build hopefulness and subsequently reduce perceived stress. Early BATA sessions emphasize greater understanding of one’s personal values to inform later behavioral change, while early MBCT sessions emphasize early awareness of one’s thoughts, feelings, and body sensations, along with a shift from task-oriented “doing mind” to a more accepting “being mind.” We conclude that individuals who reported higher perceived stress at the start of treatment benefitted from this therapy component, as evidenced by their reduced SHAPS scores at a subsequent mid-treatment timepoint (Week 4). These results suggest that changes in stressors – whether perceived or objective – are important to allow for downstream changes in hedonic functioning and reduced anhedonia.

Later in psychotherapy treatment, cross-lagged associations indicated that lower levels of perceived stress at Week 8 were predictive of lower levels of anhedonia at Week 12. We conclude that a lower level of perceived stress may be important to fully engage in the primary mechanisms of both treatments. By mid-treatment, BATA emphasizes engagement of goal-directed behavior as well as greater savoring of experience. It is possible that higher perceived stress makes disengagement from avoidance behaviors more difficult, resulting in less ability to complete valued activities. Likewise, mid-treatment MBCT includes elements of awareness of thoughts and emotions, emphasizing acceptance of difficult emotional experiences. Higher perceived stress may make acceptance more challenging. An individual who has mastered early treatment techniques appears to see the greatest improvement in later treatment weeks, as evidenced by reduced anhedonia at Week 12. Furthermore, we did not find evidence of cross-lagged paths between perceived stress and anhedonia between weeks 12 and 15 of treatment. One explanation for this lack of effect is that the association between these constructs weakens toward the end of therapy, wherein stress may become less predictive of anhedonia severity over treatment and other therapy skills contribute to reductions in anhedonia.

It is worth mentioning that aspects of behavioral activation may serve to reduce stress, at least indirectly. In BATA, participants work with their therapist to create a range of activities specific to their goal of overcoming behavioral avoidance. Some behavioral activation activities include ensuring participants achieve adequate sleep, maintaining routines, exercising, increasing meaningful social connection, work- and personal-life organization (e.g., creating and following through with to-do lists), and other self-care activities (e.g., taking a break during the day) may lead to reductions in perceived stress. Ultimately though, without testing these activities in more detail, we cannot conclude that these activities or other non-specific therapy factors were the mechanisms of early change in either intervention.

Overall, our longitudinal models suggest that perceived stress is predictive of anhedonia during psychotherapy treatment. Furthermore, the association between perceived stress and anhedonia differs in early and late treatment. These differing positive and negative effects of perceived stress on anhedonia throughout treatment require replication. Alternative explanations for this decline could include regression to the mean or poor retention leading to reduced power in the model to detect effects later in treatment (i.e., dropout).

### Limitations and Future Directions

An important limitation of the present study is that the cross-lagged panel models were carried out with a smaller sample size than is typical for structural equation models. Future studies should seek to test similar models in larger samples. Moreover, 17% (15 participants) of our sample did not complete the recommended length of psychotherapy treatment, contributing to data missingness. Together, these limitations underscore the need for replication. The current analyses also relied on self-report measures of perceived stress and anhedonia that were captured every 4 weeks of treatment. Future studies may consider exploring the association between perceived stress and anhedonia at more frequent assessments (e.g., weekly), to better capture the temporal dynamics between these constructs. Lastly, more research on the relations between perceived stress and anhedonia over time are needed to delineate the temporal precedence of the impact of stress on anhedonia.

## Conclusions

We found that individuals receiving psychotherapy treatment for anhedonia demonstrated reductions in anhedonia symptoms that were dependent upon earlier levels of perceived stress. Longitudinal models illustrated that individuals with relatively high perceived stress at the start of treatment were likely to report relatively lower anhedonia a few weeks into treatment. Later in treatment, individuals with low perceived stress were more likely to report lower anhedonia towards the end of treatment. Early treatment components are thought to reduce perceived stress, allowing for mid-to-late treatment components to exert their direct effects on anhedonia. Overall, these results suggest that perceived stress is predictive of improved symptoms of anhedonia over time in adults with transdiagnostic anhedonia. The findings presented here demonstrate the importance of including stress-reducing components in cognitive-behavioral-based anhedonia treatments.

## Data Availability

Data are available at through The National Institute of Mental Health Data Archive (NDA) collection #2595.

## Figures & Tables

**Supplementary Table 1.** Results of the autoregressive cross-lagged model (CLPM) analysis including covariates (*n*=87). Estimates are standardized coefficients. S.E. represents standard error. Tx = treatment. \*\**p*< 0.001, \**p*<.05, ^*p*<.10.

|  | Estimate | S.E. | p-value |
| --- | --- | --- | --- |
| **Perceived Stress Tx Week 15** |  |  |  |
| PSS Wk 12 | 0.787 | 0.073 | 0.000** |
| SHAPS Wk 12 | -0.140 | 0.131 | 0.283 |
| Age | 0.095 | 0.118 | 0.420 |
| Sex | 0.082 | 0.114 | 0.470 |
| Tx Group | -0.120 | 0.115 | 0.300 |
| **Anhedonia Tx Week 15** |  |  |  |
| PSS Wk 12 | 0.090 | 0.113 | 0.425 |
| SHAPS Wk 12 | 0.765 | 0.073 | 0.000** |
| Age | 0.075 | 0.113 | 0.509 |
| Sex | 0.052 | 0.107 | 0.629 |
| Tx Group | 0.090 | 0.112 | 0.423 |
| **Perceived Stress Tx Week 12** |  |  |  |
| PSS Wk 8 | 0.711 | 0.074 | 0.000** |
| SHAPS Wk 8 | 0.015 | 0.109 | 0.890 |
| **Anhedonia Tx Week 12** |  |  |  |
| PSS Wk 8 | 0.193 | 0.074 | 0.009* |
| SHAPS Wk 8 | 0.799 | 0.050 | 0.000** |
| **Perceived Stress Tx Week 8** |  |  |  |
| PSS Wk 4 | 0.434 | 0.104 | 0.000** |
| SHAPS Wk 4 | 0.181 | 0.109 | 0.097 |
| **Anhedonia Tx Week 8** |  |  |  |
| PSS Wk 4 | -0.147 | 0.103 | 0.154 |
| SHAPS Wk 4 | 0.642 | 0.072 | 0.000** |
| **Perceived Stress Tx Week 4** |  |  |  |
| PSS Wk 1 | 0.476 | 0.089 | 0.000** |
| SHAPS Wk 1 | -0.140 | 0.098 | 0.152 |
| **Anhedonia Tx Week 4** |  |  |  |
| PSS Wk 1 | -0.235 | 0.091 | 0.010* |
| SHAPS Wk 1 | 0.633 | 0.076 | 0.000** |

**Supplementary Fig 1.**
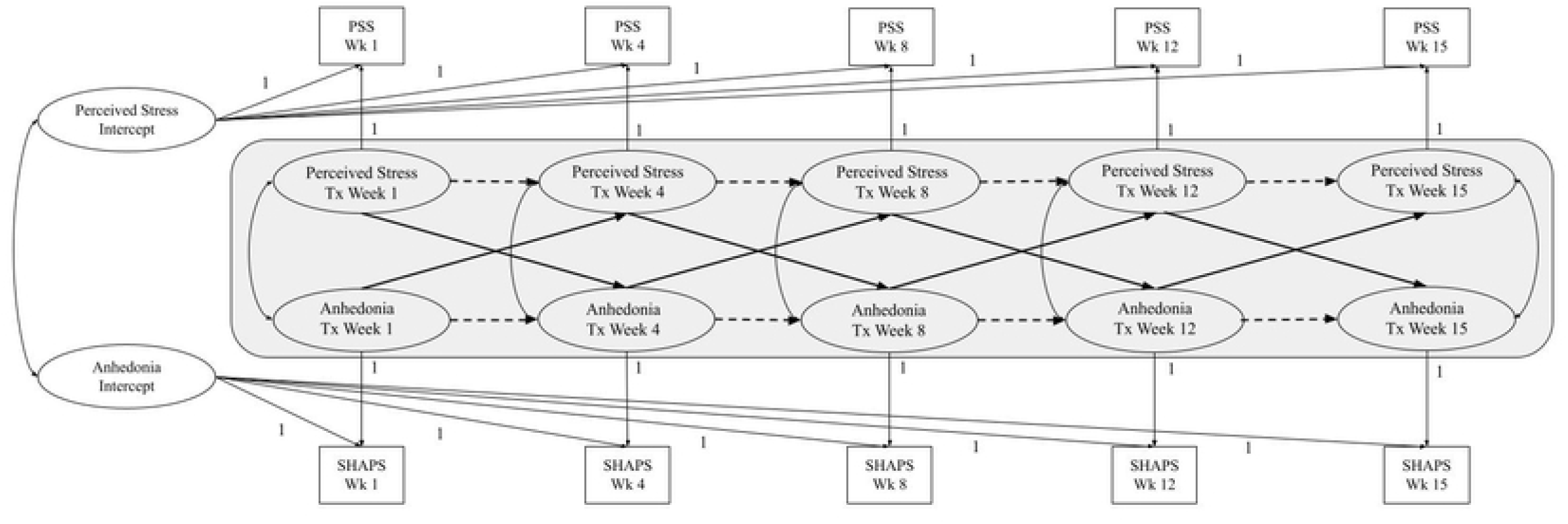
Random intercept autoregressive cross-lagged model (RI-CLPM) of anhedonia and perceived stress during psychotherapy treatment. One-headed dash lines represent auto-regressive paths; one-headed solid lines represent cross-lag paths; two-headed solid lines represent concurrent associations among variables at the same week in treatment. Variables within the gray box represent the within-person components of the model. Random intercepts represent the between-person components of the model. Path estimates and error variance terms are not included in the visualization, given that this model was uninterpretable.

